# Prediction of 7-year’s Conversion from Subjective Cognitive Decline to Mild Cognitive Impairment

**DOI:** 10.1101/2020.05.04.20091256

**Authors:** Ling Yue, Dan Hu, Han Zhang, Junhao Wen, Ye Wu, Wei Li, Lin Sun, Xia Li, Jinghua Wang, Guanjun Li, Tao Wang, Dinggang Shen, Shifu Xiao

## Abstract

Subjective cognitive decline (SCD) is a high-risk yet less understood status years before Alzheimer’s disease (AD). This work included 76 SCD individuals with two (baseline and seven years later) neuropsychological evaluations and a baseline T1-MRI. A machine learning-based model was trained based on 198 baseline neuroimage features and a battery of 25 clinical measurements to discriminate 24 progressive SCDs from 52 stable SCDs. The SCD progression was satisfactorily predicted with combined features. A history of stroke, a low education level, a low baseline MoCA score, a shrunk left amygdala, and enlarged white matter at the banks of the right superior temporal sulcus favor the progression. This is to date the largest retrospective study of SCD-to-MCI conversion with the longest follow-up, suggesting predictable far-future cognitive decline for the risky populations with baseline measures only. These findings provide valuable knowledge to the future neuropathological studies of AD in its prodromal phase.

## Introduction

Alzheimer’s disease (AD) is a neurodegenerative disorder with a slow and lengthy progression. The neuropathological process of AD is initiated at least 15-20 years before the first symptom of cognitive impairment(Elliott, Masliah, Ryan, & Silverberg, 2018; Tondelli et al., 2012). Current consensus has emphasized the need for early detection of AD based on neuroimaging (PET and MRI) (Guo, Landau, & Jagust, 2020) or invasively acquired CSF biomarkers(Frisoni et al., 2017). Detection of amnestic mild cognitive impairment (MCI), a prodromal stage of AD, maybe still too late for early intervention as the massive neuron loss and irreversible cognitive impairment may have already incurred at this stage(Ronald C. Petersen). It is crucial to explore early markers to predict possible AD conversion at an even earlier stage. Subjective cognitive decline (SCD), defined as a subjectively experienced decline in cognitive capacities in the absence of objectively measurable neuropsychological deficits, is regarded as the first indicator of AD years before MCI(Tondelli et al., 2012). Recently, the National Institute on Aging and Alzheimer’s Association (NIA-AA) updated the research guideline for AD and defined SCD as a stage 2 AD between asymptomatic (stage 1) and symptomatic MCI (stage 3)(Elliott et al.).

Accumulating epidemiologic studies support the inclusion of SCD as a pre-MCI stage and a high-risk cohort of future progression(Buckley et al., 2016; Frank Jessen et al., 2020; Mitchell, Beaumont, Ferguson, Yadegarfar, & Stubbs, 2014; Reisberg, Shulman, Torossian, Leng, & Zhu, 2010; Snitz et al., 2018). A meta-analysis study showed that the annual conversion rate from SCD to MCI is 6.6%(Mitchell et al., 2014). However, SCD can manifest in the healthy elderly or people with other disease as well. Some studies regarded subjective cognitive complaints as a benign symptom that may not lead to severe consequences(Hessen et al., 2017), while others reported that a small part of SCD might develop to cerebrovascular disease, Parkinson’s Disease, or non-Alzheimer degenerative dementia(Frank Jessen et al., 2020). The evidence altogether indicates that SCD could be a heterogeneous cohort, including those who will eventually become AD (progressive SCD or pSCD), stay stable (sSCD), and show related symptoms to other diseases. It is extremely challenging to differentiate pSCD from sSCD at the baseline. PET or CSF can detect β-amyloid or tau-related biomarkers for early AD diagnosis, but both are invasive and expensive, making it unsuitable for large-cohort screening(Elliott et al., 2018). Structural MRI (sMRI) is a non-invasive brain imaging technique that can detect AD-related morphological alterations (e.g., medial temporal lobe atrophy) in the subjects with SCD (Cherbuin, Sargent-Cox, Easteal, Sachdev, & Anstey, 2015; Dubois et al., 2018; F. Jessen et al., 2006; Meiberth et al., 2015; Peter et al., 2014; Stewart et al., 2011; Striepens et al., 2010; Tijms et al., 2018; Verfaillie et al., 2016; Yue et al., 2018). However, such findings are not conclusive for SCD compared to those for AD and MCI, possibly due to the confounding effects in the cross-sectional design used in most of the studies(F. Jessen et al., 2006; Meiberth et al., 2015; Striepens et al., 2010; Yue et al., 2018). Only a handful of longitudinal sMRI studies with only 2-5 years follow-up but such a short follow-up, which is, however, still insufficient for progression prediction (Cherbuin et al., 2015; Dubois et al., 2018; EY et al., 2019; Peter et al., 2014; Stewart et al., 2011; Tijms et al., 2018; Verfaillie et al., 2016). Identifying measurable and objective baseline markers from the SCD subjects based on non-invasive neuroimaging with a longer follow-up time for individualized early AD prediction is of great clinical significance.

Besides neuroimaging markers, clinical and demographic features are also considered to have strong predictive abilities(Livingston et al., 2017). However, none of them has been investigated in any SCD progression study, and whether they are strong enough to independently differentiate pSCD from sSCD at the baseline is yet unclear. Different sMRI indicators and clinical/demographic features may likely have complex relationships, and they could be jointly used to predict SCD-to-MCI progression. Hence, in this study, we aimed to use machine learning, an advanced multivariate pattern analysis method, to not only detect the progression-related markers jointly and objectively but also conduct an individualized differentiation between pSCDs and sSCDs. To validate our model and further test the efficacy of the identified contributive markers, we further compared the detected markers among various normal control (NC)/SCD/MCI groups with independent datasets.

## Results

### Clinical information

The demographic and clinical characteristics of the 76 SCD subjects are presented in Table 1. There are 24 (31.5%) SCD subjects showing clinical progression to amnestic MCI. Compared to sSCD, the pSCDs were less educated (7.17±4. 15 vs 9.50±3.06 years, *p* = 0.019) but with comparable baseline MoCA and MMSE scores. The two groups matched on age, gender, BMI, the status of physical diseases, lifestyle and GDS score (Table 1). At the follow-up that is seven years after the baseline scan, pSCDs developed to amnestic MCI with significant lower MoCA and MMSE scores than sSCDs but still comparable physical disease statuses and GDS score.

**Table 1.** Demographic and clinical data of the subjects with stable and progressive subjective cognitive decline.

|  | sSCD (N = 52) | pSCD (N = 24) | *t*/*λ*/*F* | *p* |
| --- | --- | --- | --- | --- |
| Age, year | 68.56 (7.19) | 71.29 (6.55) | 2.505 | 0.118 |
| Gender (male/female) | 25/27 | 11/13 | 0.033 | 0.856 |
| Education, year | 9.50 (3.06) | 7.17 (4.15) | 7.584 | 0.019 |
| Follow-up interval, month | 83.97 (0.97) | 84.16 (1.08) | 0.578 | 0.450 |
| **Baseline** |  |  |  |  |
| MoCA* | 24.04 (3.90) | 22.96 (4.49) | 0.502 | 0.481 |
| MMSE* | 27.71 (2.15) | 26.75 (2.61) | 0.038 | 0.846 |
| BMI | 24.29 (2.91) | 23.59 (4.29) | 0.725 | 0.473 |
| Smoke, year | 12.62 (17.81) | 6.83 (15.78) | 1.424 | 0.161 |
| Drink, year | 5.54 (11.71) | 7.21 (16.54) | -0.505 | 0.615 |
| Hypertension, year | 6.45 (9.37) | 7.79 (9.46) | -0.578 | 0.565 |
| diabetes mellitus, year | 1.84 (7.38) | 2.29 (6.38) | -0.260 | 0.795 |
| Hyperlipidemia, year | 1.37 (4.68) | 1.96 (5.41) | -0.489 | 0.627 |
| Heart disease (yes/no) | 16/36 | 8/16 | 0.050 | 0.823 |
| Stroke history (yes/no) | 3/49 | 4/20 | 1.211 | 0.271 |
| Surgical history (yes/no) | 29/21 | 10/14 | 1.736 | 0.188 |
| Sleep disorder, year | 1.04 (4.52) | 2.75 (7.02) | -1.094 | 0.282 |
| GDS score | 3.37 (3.90) | 2.29 (2.40) | 1.241 | 0.218 |
| Social support score | 36.52 (9.59) | 33.42 (10.67) | 1.265 | 0.210 |
| **7-year follow-up** |  |  |  |  |
| MoCA* | 23.12 (3.54) | 17.33 (3.82) | 29.358 | <0.001 |
| MMSE* | 27.58 (1.95) | 24.08 (3.87) | 17.497 | <0.001 |
| BMI | 25.01 (2.99) | 23.69 (3.95) | 1.607 | 0.112 |
| Sleep disorder, year | 0.96 (2.20) | 2.25 (4.50) | -1.336 | 0.192 |
| Hearing loss, year | 0.43 (1.10) | 0.79 (1.44) | -1.093 | 0.282 |
| GDS score | 4.52 (3.47) | 5.29 (4.67) | -0.806 | 0.423 |
| Hypertension, year<sup>†</sup> | 0.06 (0.31) | 0.25 (0.74) | -1.229 | 0.230 |
| Diabetes mellitus, year<sup>†</sup> | 0.500 (1.49) | 0.46 (1.25) | 0.107 | 0.915 |
| Hyperlipidemia, year<sup>†</sup> | 0.21 (0.87) | 0.08 (0.41) | 0.685 | 0.495 |
| Heart disease (yes/no)<sup>†</sup> | 6/46 | 4/20 | 0.062 | 0.803 |
| Stroke history (yes/no)<sup>†</sup> | 7/45 | 2/22 | 0.068 | 0.794 |
| Surgical history (yes/no)<sup>†</sup> | 9/43 | 8/16 | 2.429 | 0.119 |

Data presented are the means (standard deviations) or sample size (N). sSCD, stable subjective cognitive decline, pSCD, progressive subjective cognitive decline, MoCA, Montreal Cognitive Assessment, MMSE, Mini-Mental State Examination, BMI, Body Mass Index, GDS, Geriatric Depression Scale.
\*All the analyses of neuropsychological variables (i.e., MoCA and MMSE) were conducted after controlling for age, gender, and education.
<sup>†</sup> Newly diagnosed after the baseline during the 7-year follow-up.

### Machine learning-based pSCD vs. sSCD classification

With 233 clinical and MRI features, cost-sensitive support vector machines (CSVM) achieved a satisfactory pSCD vs. sSCD classification performance (accuracy, 69.74%, sensitivity, 62.50%, specificity, 73.08%, F1 score, 0.5660). Specifically, the algorithm successfully identified 15 pSCDs out of 24, and 38 sSCDs out of 52. Supplementary Table 1 and Supplementary Table 2 lists all the features and the confusion matrix.

### The most contributive features in SCD progression prediction

Five features were identified and consistently selected as contributive features in pSCD vs. sSCD classification (see details of the selection frequency in Supplementary Table 1). They are baseline stroke history, years of education, baseline MoCA score, baseline volume of the left amygdala, and baseline WM volume at the banks of the right superior temporal sulcus (wmSTSbanks). Figure 1A and Supplementary Table 3 show the group comparison results for these five features between pSCDs and sSCDs. In addition to education level, both MRI-based features (see Supplementary Figure 1 for locations) show significant group differences (*p* < 0.05, uncorrected). The pSCD individuals show lower MoCA scores at baseline, more likely to have stroke history, lower education level, decreased left amygdala volume, and increased right wmSTSbanks volume, compared to sSCDs. With the five contributive features, we derived a clinically feasible, much simpler, automatic prediction model for SCD progression based only on three clinical/demographic features and two MRI features at the baseline. Figure 1B shows the ROC curve of these five contributive features with a refined SCD progression prediction model and the AUC reached 0.7997. The distributions of the decision scores obtained by the scoring function learned by this prediction model for all the subjects are shown in Figure 1C, indicating a satisfactory separation of the two groups with only a few baseline features.

**Figure 1.**
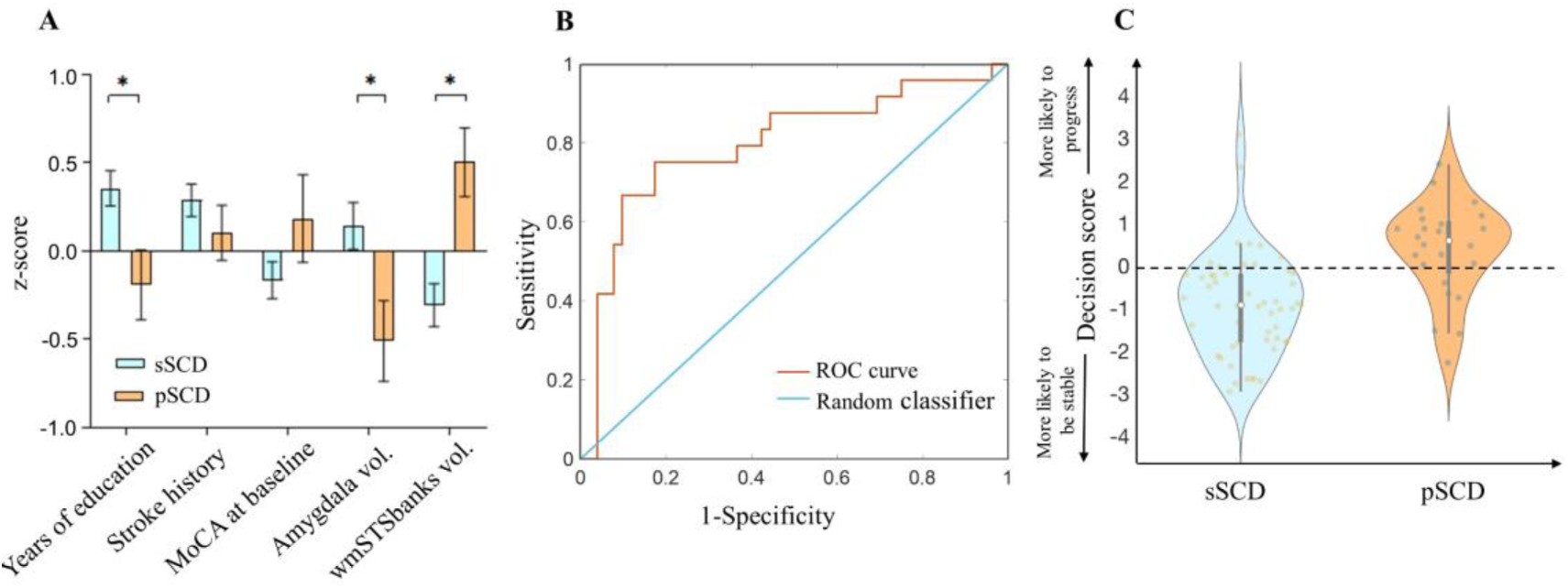
The five most contributive features. (A) The error bar plots of the five most contributive features in z-scores identified by cost-sensitive support vector machines (CSVMs) in the sSCD vs. pSCD classification. Error bars indicate standard errors. (B) The ROC curve generated by using the five most contributive features to classify pSCDs and sSCDs (AUC = 0.7997). (C) The decision scores generated by the refined SCD progression prediction model with the five features. The white dot in the middle is the median value and the thick black bar in the center represents the interquartile range. The thin black line extended from it represents the upper (max) and lower (min) adjacent values in the data. sSCD, stable subjective cognitive decline, pSCD, progressive subjective cognitive decline, wmSTSbanks, baseline white matter volume at the banks of the right superior temporal sulcus, ROC, receiver operating characteristic, AUC, area under the curve, MoCA, Montreal Cognitive Assessment.

### Relationship between MRI features and cognitive score

The partial correlation analysis between the two important MRI features (volume of the left amygdala and the right wmSTSbanks) and the MoCA scores (baseline, follow-up, and the changes), after controlling education, stroke history, age, and gender, revealed a negative association between the baseline left amygdala volume and the follow-up MoCA score (r = −0.636, *p* = 0.003, significant after Bonferroni corrections) for the pSCD subjects (Figure 2A). No significant association was found for the sSCD subjects. No correlation was found between the baseline volume of the right wmSTSbanks and the baseline MoCA scores or its changes.

**Figure 2.**
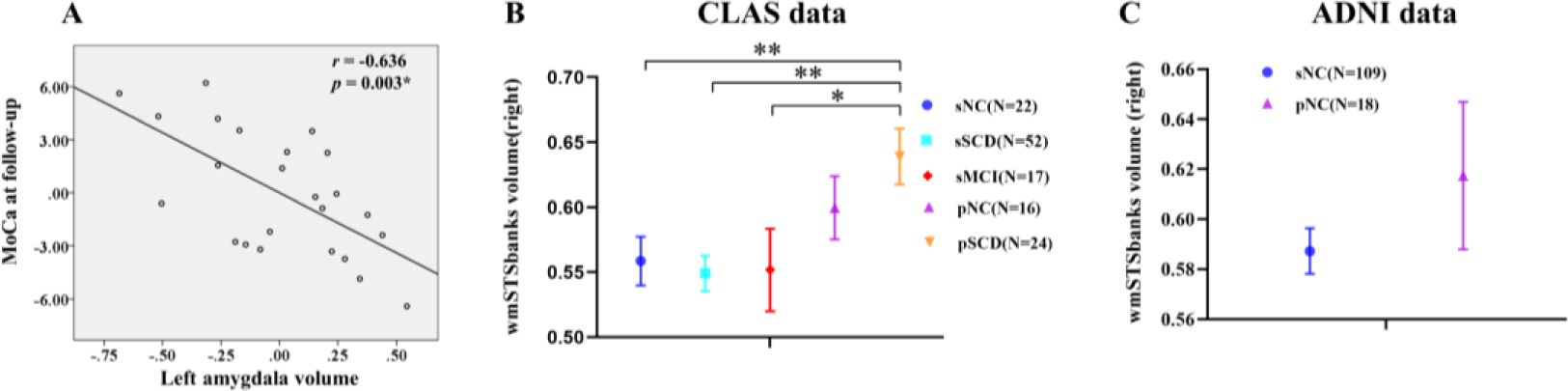
Clinical correlation, and validations based on CLAS study and ADNI. (A) The scatter map shows a significant correlation (after removing the confounding effects) between the volume of the left amygdala and the MoCA score at the follow-up in the pSCD group (p < 0.05, after Bonferroni correction, where the uncorrected p threshold equals 0.05/(2 groups × 2 MRI features × 3 MoCA scores), or 0.0042). (B) Comparison of the baseline right wmSTSbanks’ volume among the groups with different cognitive functions at the baseline and the follow-up from the CLAS database, with additional cohorts of sNC, pNC, and sMCI compared to pSCD, separately (* indicates p < 0.1, after Bonferroni correction with a threshold of 0.1/4 = 0.025 (as four different groups were compared with pSCD) to the original p-values; ** indicates p < 0.05, after Bonferroni correction with a threshold of 0.05/4 = 0.0125 to the original p-values). (C) Comparison of the baseline right wmSTSbanks volume between sNC and pNC groups from the ADNI data. Error bars indicate standard errors. The region volume was standardized (no unit). sNC, stable normal control, sSCD, stable subjective cognitive decline, pNC, progressive normal control, pSCD, progressive subjective cognitive decline, sMCI, stable mild cognitive impairment, MoCA, Montreal Cognitive Assessment.

### Structural connections of the right wmSTSbanks

For possible WM connections to the right wmSTSbanks (i.e., the only WM feature from the MRI detected by the machine learning-based pSCD vs. sSCD differentiation). dMRI tractography with an independent dMRI data from a healthy elderly cohort (Supplementary Experiment 1) showed that there are many WM fibers could pass through the right wmSTSbanks and linking to the right amygdala and the right hippocampus, two regions targeted by the AD pathology (Supplementary Figure 2). In addition, there are many other cortical regions, especially the right middle, superior, and inferior temporal gyri, as well as the right interior parietal and supramarginal areas and right putamen, could be reached by the WM fibers that passing through the right wmSTSbanks (Supplementary Figure 3).

### Validation based on independent data sets

From the CLAS database, we further identified sNC, pNC, sMCI subjects and extracted the same features from them. Together with the sSCD, all four groups were compared with the pSCD group, separately, with two-sample *t*-tests with Bonferroni correction. We found a similar trend that sNC, sSCD, and sMCI, whom were all potentially not affected by AD pathology, tend to have lower baseline wmSTSbanks volume compared with pSCD, with the pNC specifically sitting in-between the three stable groups and the sSCD group. Specifically, we found a significant difference (*p*_corrected_ < 0.05) between sNC and pSCD in addition to the significant difference between sSCD and pSCD, as well as a trend-to-significant difference (*p*_corrected_ < 0.1) between sMCI and pSCD (Figure 2B, see details in Supplementary Experiment 2). Furthermore, the combined progressive group (pSCD+pNC) also showed a greater baseline right wmSTSbanks volume than the combined stable group (sSCD+sNC) (Supplementary Figure 4). From the ADNI data, although no significant difference was found from the comparison of the baseline volume of the right wmSTSbanks between sNCs and pNCs (*p* = 0.24) due to lower statistical power caused by small sample size (due to short follow-up), we still spotted a similar trend to the main results, i.e., pNCs had a higher right wmSTSbanks volume than the sNCs (0.62±0.13 vs. 0.59±0.09, Figure 2C, see details in Supplementary Experiment 3).

## Discussion

SCD is recognized as a high-risk status of AD with a normal cognitive level, which is earlier than the stage of MCI(Buckley et al., 2016; Mitchell et al., 2014; Wolfsgruber et al., 2015). In the current study, instead of predicting MCI progression, we focus on the detection of possible AD at an even earlier stage, i.e., predicting whether SCD converts to MCI. We were able to do so because of the unique CLAS dataset that focused on the risky population at even earlier stage compared to MCI and had an extremely long follow-up time (seven years). Based on a well-designed machine learning model with both comprehensive clinical features and MRI features, we demonstrated that the SCD individuals who would develop to amnestic MCI later might be identified and differentiated from the sSCDs with a combination of a few clinical information, psychometric scores, and an easily obtained, baseline grey-matter and white-matter volumetric data based on sMRI. Our results indicated that incident stroke, fewer years of education, lower baseline MoCA score, smaller left amygdala, and larger white matter at the banks of right superior temporal sulcus jointly favored amnestic MCI progression. We also showed that with a state-of-the-art, data-driven feature selection and classification, the sSCD could be individually separated from the pSCD in an automatic and objective way with satisfactory accuracy. We then proposed a clinically feasible, much-simplified prediction model with only five (three clinical and two sMRI-derived) baseline features with which an AUC of 0.8 was reached. The strengths of this study include an unprecedently long-term follow-up, a targeted risk population (SCD) from well-characterized community samples, an advanced multivariate pattern recognition algorithm that jointly identified the associations among the clinical and brain MRI features towards an effective feature set, and a demonstrated feasibility of early computer-aided diagnosis of pSCD individuals.

While it is important to identify which individuals with SCD eventually develop clinically significant cognitive impairment years later, long term follow-up is crucial because it might take almost a decade for a subject with “compensatory normal cognition” to change from SCD to MCI (Molinuevo et al., 2017), especially for community-based cohorts(Snitz et al., 2018). However, to our best knowledge, only four SCD follow-up studies had investigated the link between baseline brain biomarkers and incident clinical progression, but all of them only followed up for 2-5 years(Dubois et al., 2018; EY et al., 2019; Tijms et al., 2018; Verfaillie et al., 2016). This might not be sufficient to fully investigate the conversion from SCD to MCI. One study only identified four out of 318 SCD individuals who progressed to prodromal AD within 30 months(Dubois et al., 2018), while another study reported that community-based SCD subjects confer much lower risk of progression to MCI over three years compared to clinical cohorts(Snitz et al., 2018). Not only cutting off sample sizes, insufficient follow-up time could also misidentify many SCDs who could potentially convert to MCI, which could result in a highly mixed and heterogenous sSCD group, further reducing statistical power. Furthermore, compared to the subject recruitment from clinical sites, those recruited from communities can empower the AD early detection by enabling even earlier detection, even for those who have not developed cognitive impairment yet (on contrast, clinical cohorts could have more or less developed symptoms already at the baseline). Our study is an SCD conversion study with a long (> five years) follow-up, with advanced feature searching strategies from comprehensively engineered features to identify pivotal factors that could provide early predictive value.

Our most important finding is the early enlargement of the right wmSTSbanks in the pSCD vs. sSCD. This feature characterizes the relative WM volume beneath the cortical part of the superior temporal sulcus (STS). STS is a “chameleon” in the human brain and a high-order association cortex that receives connections from different sensory modalities(Hein & Knight, 2008). It is believed to be involved in diverse cognitive functions, such as audiovisual integration, as well as motion, speech, and face processing(Hein & Knight, 2008). The WM under the STS may play a crucial role in efficient information transmission to support the multifunctionality of the STS. This is further proved by our fiber tracking result, which shows dense fiber connections with lateral and medial temporal cortices, an area for multimodal information integration and the language- and memory-related cognitive functions(R, MA, S, & G, 2011). Meanwhile, STS was consistently reported to be affected by AD neuropathology (e.g., neurofibrillary tangles and neuronal loss) in a very early stage. The neural loss in the STS was found to be correlated with duration of illness, ranging from no measurable loss in the early stage (with a duration of less than one year) to more than 75% neuron reductions in the severe AD stage(Gomez-Isla et al., 1997). Hence, we interpret our finding of early increment in the STS-related WM volume as a result from increased axonal connections as compensation of other affected regions to preserve normative cognitive ability in the SCDs who would convert to AD and such compensation could be enabled by the largely preserved STS neurons working with an elevated effort or load in the start-up phase of clinical AD. Our additional comparisons with sNC, sMCI, and pNC cohorts from the CLAS database further validate such a compensation hypothesis (Figure 2B). Of note, additional support came from the results with a similar trend of increased baseline wmSTSbanks volume in pNC compared to sNC from the ADNI data (Figure 2C), which might become significant providing sufficient follow-up time and more balanced sample size. From the functional study point of view, an 18FDG-PET study shows increased metabolism in the right WM adjacent with the inferior parietal lobe in SCD subjects compared to NCs(Scheef et al., 2012). Another study revealed increased regional cerebral blood flow (rCBF) along the default mode network regions in SCD compared to NCs(Wang et al., 2013). These areas are either closed to or include the wmSTSbanks, further supporting our compensation hypothesis.

Another potential reason for the greater WM underneath the STS could be grey-matter atrophy in the adjacent areas. A previous study reported decreased grey matter at the banks of STS in the MCI subjects who converted to AD within three years(Killiany et al., 2000). A very recent PET study with ADNI data also found that cognitively normal individuals with high Aβ burden in the cortical region of the banks of STS were at an increased risk of cognitive decline(Guo et al., 2020). Of note, we used a ratio of wmSTSbanks volume over the total white matter volume as it is a conventional way to exclude confounding effects (e.g., global WM atrophy)(Nordenskjold et al., 2013), there could be a possibility that it is the shrunk total white matter volume that led to such a result. However, we found that it is less likely because we found that the pSCD still showed larger volume than sSCD even without conducting such a global normalization (Suppliementary table 4).

Another contributive sMRI feature is the decreased baseline volume of the left amygdala in pSCD compared to sSCD. Amygdala atrophy in SCD was consistently reported(Hu et al., 2019; Striepens et al., 2010; Yue et al., 2018), especially by a recent study that revealed a smaller left amygdala in the SCDs with CSF biomarker (Aβ42) compared to those without(Hu et al., 2019). Another long-term follow-up study also suggests that atrophy of the amygdala in cognitively intact elderly people could predict dementia occurred 6 years later.(den Heijer et al., 2006) Also, from the cognitively intact elderly cohort, Tondelli et al. reported the reduced amygdala volume was detectable at least four years before any cognitive symptoms.(Tondelli et al., 2012) Moreover, decreased amygdala is also be associated with TDP43-related dementia(N et al., 2019). Hence, atrophy in this area in SCD may be an early marker for future cognitive decline. It is noted that we only found a relatively weak statistical difference (p = 0.011) in the amygdala by statistical analysis, which means that this region could be undetectable from a large number of candidate features if using traditional mass-univariate analysis. Instead, with advanced feature selection and multivariate pattern analysis in a unified framework of machine learning, we were able to detect such a contributive sMRI feature together with others.

While the pSCD subjects had a smaller baseline amygdala compared to sSCDs, we found a negative correlation between the baseline amygdala volume and the MoCA scores at the follow-up across the pSCD subjects while no such correlation was found from sSCD. This indicates that, for pSCD, larger amygdala could be associated with worse cognitive outcome. Such results seem contradictory as positive correlation was originally anticipated as sSCD (with good outcomes) was found to have a larger amygdala compared to pSCD (but we found that a larger amygdala in pSCD predicted worse outcome). Therefore, we think that the negative correlation could support a well-known hypothesis that higher cognitive reservation (as reflected by a larger amygdala at the baseline) is associated with a more rapid decline later (and therefore has much worse cognitive outcomes afterward) for the risky populations(Amieva et al., 2014). We further came into an updated hypothesis, that is, as long as the baseline amygdala size is below average, those with a larger amygdala may cover up baseline symptoms and, once they become clinically significant, the subjects could have more rapidly declined cognitive abilities. This finding might have helpful for developing possible preventative solutions or treatments in the future.

In addition to the sMRI features, the study also revealed three contributive clinical features in distinguishing the cognitive outcome of SCDs, including baseline MoCA, years of education, and history of stroke, all of which have been reported as indicators of poor cognitive prognosis in the previous studies(Amieva et al., 2014; K et al., 2019; Nasreddine et al., 2005). Again, traditional statistical analysis could not reveal significant differences in MoCA and stroke history (Supplementary Table 3). However, this does not contradict our findings, as our prediction model considered these features jointly instead of independently. Our method is quite effective, which is also because we utilized an effective feature selection algorithm using Relief, an algorithm that takes relationships among features into consideration. Furthermore, we derived a simplified model with only five features, all of which can be obtained at the baseline. This renders our model a merit of predicting SCD conversion at the baseline seven years before noticeable cognitive impairment, suitable for large-cohort screening in the future.

However, this study also has limitations. First, our study does not consist of annual follow-ups, which maybe helpful for timely identifying SCD’s conversion to MCI. Second, although we followed up on our SCD subjects for a long time compared to the previous studies, it is still likely that more SCD will progress to MCI at later time(Kaup, Nettiksimmons, LeBlanc, & Yaffe, 2015). Third, our data does not include FDG-PET, amyloid-β marker, or APOE genotype, making it unclear whether the progression was due to AD or not.

Nevertheless, this is the first sMRI study of SCD subjects with unprecedentedly long follow-up time. We, with advanced machine learning from the comprehensively and thoroughly engineered feature sets, identified key baseline early AD markers comprising both clinical and neuroimaging features with an individualized SCD progression prediction task. We further proposed a much simpler prediction model with only five features and a satisfactory predictive accuracy. Our results provide a potentially feasible and objective early diagnostic tool and help better understanding of the pathology of AD in its prodromal phase.

## Methods

### Study cohorts

The study is part of the China Longitudinal Ageing Study (CLAS) of Cognitive Impairment, which is a community-based study initiated in 2011(Xiao et al., 2016; Xiao et al., 2013). The current data constitute samples from Shanghai where all the subjects received a baseline T1-MRI scan. Some of the data was used in a cross-sectional study on SCD(Yue et al., 2018). In the current study, as the 7-year follow-up has recently finished, we could include all the subjects with both baseline SCD diagnosis and the follow-up visit. Of all the 111 SCD subjects defined at the baseline, the 7-year follow-up was completed in 92 (82.8%) subjects (Table 1). The SCDs with and without follow-up did not differ in gender (male/female: 44/48 vs. 9/10), age (69.23±7.49 vs. 69.84±6.90 years), years of education (9.01±3.78 vs. 9.21±4.44 years), the Mini-Mental State Examination (MMSE) score (27.55±2.23 vs. 27.16±2.59), or the Montreal Cognitive Assessment (MoCA) score (23.54±4.56 vs. 23.00±4.66). Of note, even the 7-year follow-up cannot guarantee the conversion from SCD to AD; therefore, we focused on on predicting SCD conversion to amnestic MCI (the pSCD is hereby defined accordingly from here on). Although we cannot guarantee that all the converted cases were due to AD, we still remove 16 SCD subjects diagnosed with follow-up evaluations as other neurodegeneration diseases, psychogenic diseases, or organic etiologies to avoid confounding effects. Figure 3 shows a flowchart of participant selection. The study protocol was approved by the Ethical Committee of the Shanghai Mental Health Center. All participants signed written informed consent before enrollment.

**Figure 3.**
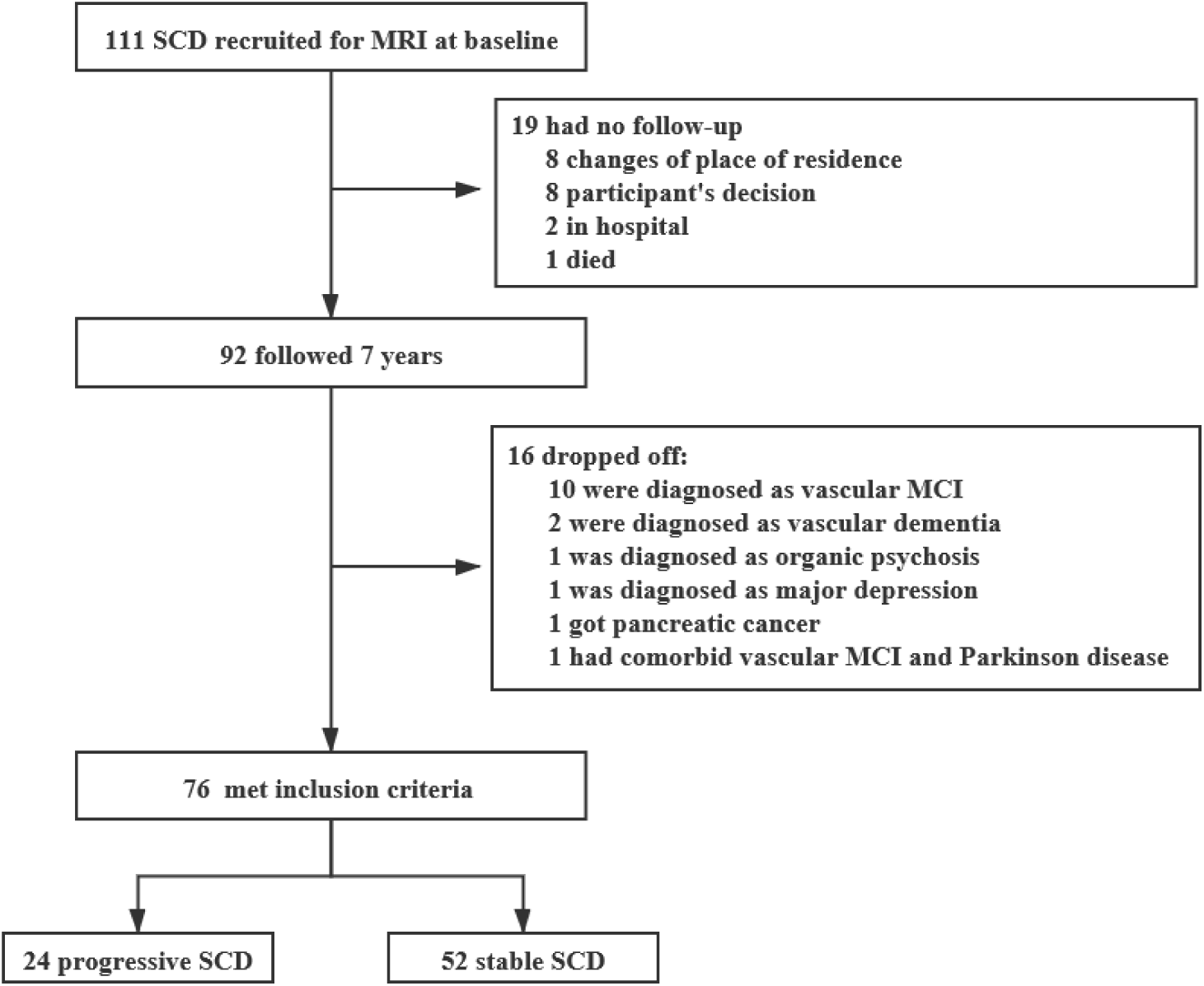
Subject selection flowchart. SCD, subjective cognitive decline, MCI, mild cognitive impairment.

All the SCD participants were assessed by self-report and diagnosed according to the following criteria(Frank Jessen et al., 2014; Molinuevo et al., 2017): 1) the onset age of memory decline is > 60 years old; 2) presence of gradual memory decline has persisted for ≥ 6 months; 3) objective memory performance at baseline is within the normal range. More details on the SCD diagnosis procedure are listed elsewhere(Yue et al., 2018). MCI was clinically diagnosed according to the Peterson’s criteria of MCI with consideration of comorbid conditions. At the follow-up visit(R. C. Petersen et al., 1999), we had 24 pSCD subjects who had progressed to MCI and the 52 sSCD subjects who kept cognitive normal, none of which converted to AD at the follow-up visit.

### Subject assessment

We performed a comprehensive battery of sociodemographic and physical health measurements for each subject. As potential risk factors of SCD progression(Livingston et al., 2017), sociodemographic data (gender, age, years of education, years of smoke, years of drink, and Body Mass Index (BMI)) and status of somatic diseases (years of hypertension, hyperlipidemia, diabetes mellitus, sleeping disorder, incident stroke, heart disease, and any type of surgery) at the baseline were collected, as well as the new onsets of the above somatic diseases and years of hearing loss acquired over the follow-up period. Three psychological assessments were carried out at the baseline, including MoCA (measuring overall cognitive performance), geriatric depression scale (GDS, also acquired at the follow-up due to the close association between depressive symptoms and cognitive performance), and social support questionnaire (measuring perceptions of social support and its satisfaction).

### MRI data acquisition and analysis

T1-weighted 3D magnetization prepared rapid gradient echo (MPRAGE) sMRI were acquired from a 3.0T MRI scanner (Siemens MAGNETOM VERIO, Germany) with the following parameters: TR = 2300 ms, TE = 2.98 ms, flip angle, 9°, matrix size, 240 × 256, field of view (FOV), 240 × 256 mm, slice thickness, 1.2 mm, and number of slices, 176. All the sMRI data were processed using Clinica (Routier et al., 2018) (www.clinica.run) in FreeSurfer v6.0 (surfer.nmr.mgh.harvard.edu), including segmentation of the subcortical structures, extraction of cortical surfaces, cortical thickness estimation, spatial registration, and parcellation into 46 global structures. Quality control was carefully conducted by overlapping the output parcellations on the FreeSurfer’s template and visual assessment was carried out to ensure the registration and parcellation quality. Besides the 46 global volumetric measurements of different brain structures, we further derive 68 cortical, 68 white-matter (WM), and 16 subcortical regions of interest (ROIs) based on the Desikan-Killiany atlas(Desikan et al., 2006). For each cortical ROI, mean cortical thickness was extracted without normalization(Westman, Aguilar, Muehlboeck, & Simmons, 2012). For each WM or subcortical ROI, the regional volumetric measure was extracted and normalized by the total volume of WM and subcortical regions, respectively. All the 198 (46 + 68 + 68 + 16) MRI features were listed in Supplementary Table 1.

### SCD progression prediction

Due to the potential redundancy in the 223 (25 clinical + 198 MRI) features, it is necessary to select the most predictive features to train a concise model and avoid overfitting. We used a three-stage feature selection scheme. First, a Relief algorithm was employed to rank the features according to the extent of associations with the labels (i.e., pSCD and sSCD), which has been widely applied in machine learning tasks to improve classification performance, e.g. genetic analysis for diseases, Parkinson’s disease diagnosis and contributive feature identification(Urbanowicz, Meeker, La Cava, Olson, & Moore, 2018). Considering contextual information during ranking, Relief can properly handle strong dependency among features. Specifically, it generates a robust feature ranking by identifying the feature value differences between nearest-neighbor instance pairs. The number of neighbors was selected from a range between 1 and 20 at a step of 1. Therefore, we generated 20 ranking lists of features. Second, an overall ranking for all the features was obtained by integrating the 20 ranking lists and calculating the occurrence frequency of each feature in the top-50 of each ranking list. Finally, the classification performance based on CSVMs, which better handles the imbalanced samples than the traditional SVM) was derived to evaluate the ranked feature set and determine the predictive features(Park, Luo, Parhi, & Netoff, 2011). F1 score (defined as the harmonic average of the precision and sensitivity, i.e., F1 score = 2 × (precision × sensitivity) / (precision + sensitivity)) was taken as the classification performance evaluation metric for further feature selection and parameter optimization of CSVM. A sequential forward selection strategy was adopted in this step, which sequentially added the features according to their ranks until adding a new feature did not improve the CSVMs’ performance. The same CSVMs were used for SCD progression prediction. We set a higher misclassification penalty (with a freely estimable cost ratio, pSCD:sSCD) for the pSCD samples compared to those for the sSCD samples in the CSVMs to alleviate the problem caused by unbalanced samples, as pSCDs were much fewer than sSCDs.

The prediction model was validated by nested leave-one-out cross-validation (LOOCV). The feature set selected by the inner LOOCV according to the above method was used to train the classifier and the final prediction performance (including accuracy, sensitivity, specificity, and F1 score) was obtained by the outer LOOCV. In each iteration of the outer LOOCV, one subject was left out as a testing sample and the remaining 75 subjects were for training (these 75 subjects were fed into the inner LOOCV where the predictive features were selected). The trained model was applied to the testing sample for generating a predicted label to compare with the ground truth. The outer LOOCV went through all the subjects. The frequency of each feature being selected across the 76 outer LOOCV iterations was used to assess feature importance. A feature was regarded as a contributive feature if this frequency is higher than 95% (i.e., selected from 72 out of 76 LOOCV iterations). Figure 4 shows the flowchart of the entire analysis.

**Figure 4.**
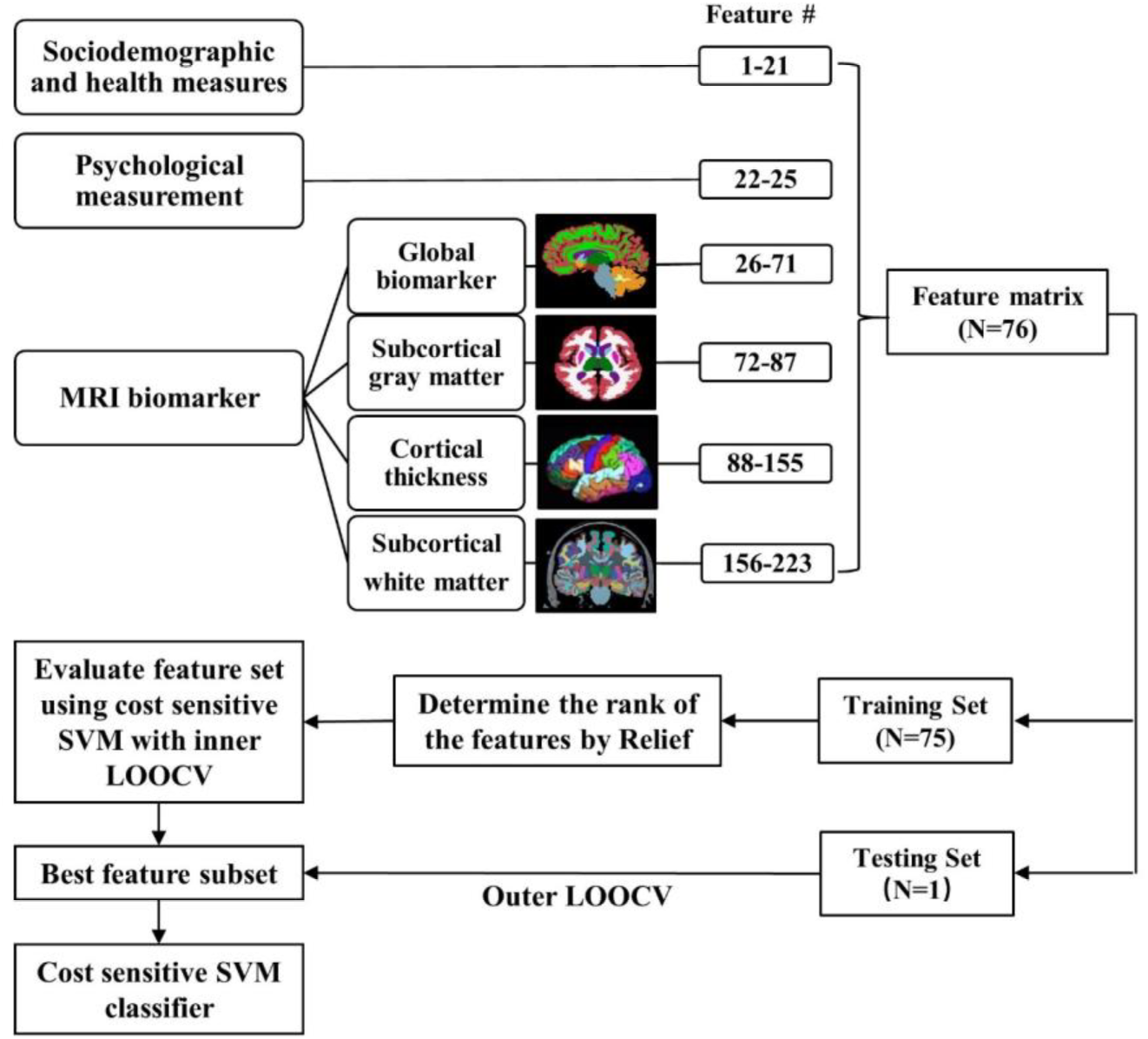
Schematic flowchart of the SCD-to-MCI prediction framework based on the features extracted from sMRI and clinical measurement. LOOCV, leave-one-out cross-validation, SVM, support vector machine, SCD, subjective cognitive decline, MCI, mild cognitive impairment.

To facilitate clinical application, after the contributive features were identified, we re-trained a refined SCD progression prediction model with only these contributive features as a representative prediction model. Based on this model, we drew a receiver operating characteristic (ROC) curve and used the area under the curve (AUC) to measure the discriminant ability in the prediction of SCD progression. We further derived an intuitive “decision score” for each subject according to the scoring function learned by such a representative prediction model.

### Statistical analysis

Group comparisons of the demographic and clinical data were conducted using two-tailed independent samples t-tests for continuous variables and *χ*^2^-tests for dichotomous variables with SPSS 19.0 (IBM Corp.). Differences in the global cognition (i.e., MoCA and MMSE) between groups were tested after controlling for age, gender and years of education. After detecting contributive features, a *t*-test or *χ*^2^-test was carried out for each of them to see if there was any group difference between the pSCD and sSCD groups at the baseline. Specifically, we evaluated whether there were unique contributions between the identified MRI features and the cognitive measurement (as evaluated by MoCA) in the pSCD group and sSCD group, separately. This was achieved by partial correlation analysis to remove the possible influence of other non-MRI contributive features (education level and stroke history) as well as age and gender. For the partial correlation, MoCA scores at the baseline and follow-up, as well as 7 years’ MoCA changes were separately used to investigate how the identified MRI features were associated with cognitive abilities (either with the two terminal scores or their changes). Bonferroni correction was conducted for multiple comparison correction.

### Result evaluations with independent datasets

In addition to the main analysis, we analyzed various independent datasets to further validate and evaluate the main findings from the CLAS data. First, for the identified contributive MRI feature at the WM (i.e., the banks of the right superior temporal sulcus), we checked whether there were fiber connections between this region and amygdala and hippocampus, two regions specifically targeted by AD, based on an independent diffusion MRI (dMRI) tractography dataset that was used in our previous study (from 15 (10 females) healthy elderly subjects with mean age of 70.6±6.2 years, see Supplementary materials)(Li et al., 2016). We investigated which cortical areas could be reached by the tractography streams passing through the right wmSTSbanks (see details on fiber tracking in Supplementary Experiment 1). Second, we checked the volume of the wmSTSbanks in different NC cohorts (including a group of stable NCs or sNC and another group of progressive NCs or pNC) and a stable MCI (sMCI) cohort, also selected from the CLAS database (but with smaller sample size compared to the sSCD/pSCD in the main analysis), to see if there was any consistent (possibly much earlier, as progressive NCs were used) trend with the main results (see details in Supplementary Experiment 2). Third, we used sNC and pNC subjects from an independent ADNI2 dataset (http://adni.loni.usc.edu/), another widely adopted longitudinal dataset for tracking AD progression to investigate if any similar trend can be found (see details in Supplementary Experiment 3). The follow-up time for the ADNI data is 45.39±7.89 months (~ 4 years), smaller than the CLAS data.

## Data Availability

Imaging and clinical data as well as the computing code used in the current manuscript are available from the corresponding authors upon request from any qualified investigator.

## Acknowledgements

This work was supported by the China Ministry of Science and Technology (2009BAI77B03), Shanghai Mental Health Center (CRC2017ZD02, 2018-FX-05), Shanghai Clinical Research Center for Mental Health (SCRC-MH, 19MC1911100), the National Natural Science Foundation of China (81830059), Shanghai Jiaotong University School of Medicine (CBXJ201815, YG2016MS38), and Shanghai Municipal Human Resources Development Program (2017BR054).

